# The Influence of Long-Term Supplementation with *Lactiplantibacillus plantarum* LP28 on NK Cell Activity and Upper Respiratory Symptoms in Middle-Aged and Older Adults: A Randomized, Double-Blind, Placebo-Controlled Trial

**DOI:** 10.64898/2026.11.08.26367495

**Authors:** Zheng-Yi Li, Mei-Hui Tseng, Chung-Min Shieh, Mao-Chia Chang, Han-Yin Hsu, Chia-Chia Lee, Hsiao-Wen Huang, Ching-Lin Wu, Jeng-Jer Shieh

**Author notes:** Zheng-Yi Li and Mei-Hui Tseng contributed equally to this work as the first authors. Correspondence to Ching-Lin Wu: Graduate Institute of Sports and Health Management, National Chung Hsing University, Taichung City, Taiwan, No. 145 Xingda Rd., South Dist., Taichung City 402, Taiwan, Correspondence to Jeng Jer Shieh: Institute of Biomedical Sciences, National Chung Hsing University, Taichung City, Taiwan No. 145 Xingda Rd., South Dist., Taichung City 402, Taiwan.

## Abstract

**Background:** Population aging and immunosenescence pose immense challenges to healthcare systems by increasing susceptibility to infections. *Lactiplantibacillus plantarum* LP28 (*L. plantarum* LP28) is a strain isolated from traditional Taiwanese dried tofu with significant immunomodulatory potential.

**Objective:** To evaluate the clinical efficacy of long-term *L. plantarum* LP28 supplementation in common cold prevention and its effect on systemic immune biomarkers in middle-aged and elderly individuals.

**Methods:** In this 12-week, randomized, double-blind, placebo-controlled trial, 102 middle-aged and older adults (aged 50–75 years) were assigned to receive an oral probiotic (5 × 10^9^ CFU *L. plantarum* LP28/capsule, two capsules daily; *n* = 51) or a placebo (*n* = 51). Cold incidence and symptom severity were evaluated by the Wisconsin Upper Respiratory Symptom Survey (WURSS). Immune cell populations, serum/salivary markers, and natural killer (NK) cell activity were quantified using flow cytometry, ELISAs and NK cell cytotoxicity assays, respectively. The data were analyzed using a refined per-protocol approach and outlier-removed sensitivity analyses to ensure robustness. Independent *t* tests, chi-square/Fisher’s exact tests, and Mann – Whitney *U* tests were used for statistical comparisons.

**Results:** The probiotic group exhibited a notable reduction in cold incidence (34.78% vs. 52.00% in the placebo group) and a lower cumulative symptom burden. In terms of immunomodulatory effects, *L. plantarum* LP28 supplementation significantly increased serum interleukin (IL)-12 levels and NK cell cytotoxicity, reinforcing innate immune defense through the activation of the IL-12/NK cell axis. This was accompanied by a significant reduction in the level of the inflammatory marker C-reactive protein (CRP).

**Conclusions:** *L. plantarum* LP28 supplementation serves as an effective supportive strategy for promoting healthy aging by increasing respiratory defense resilience and mitigating systemic inflammatory stress through pleiotropic immune modulation.

**Clinical Trial Registration:** This trial was registered at ClinicalTrials.gov under the registration number NCT06724185 (https://clinicaltrials.gov/study/NCT06724185).

## Introduction

Population aging has become a definitive global trend. Currently, the number of individuals aged 65 years and over is 727 million worldwide, accounting for 9.3% of the total population. This proportion is projected to increase steadily to 16% over the next three decades [1]. The aging process is characterized by a progressive decline in immune efficacy. Both innate and adaptive immunity naturally weaken over time—a phenomenon clinically referred to as immunosenescence. This state of immune dysfunction makes older adults more susceptible to disease, diminishes their response to vaccinations, and increases the prevalence of systemic inflammation [1]. Historical data from 1976 to 2007 in the United States indicate that approximately 88% of annual influenza-related deaths were among individuals older than 65 [2]. The decline in immunity among elderly people not only increases infection and mortality rates during seasonal influenza outbreaks but also imposes a substantial burden on healthcare systems, society, and the global economy.

The harmful effects of aging on the innate immune system include mucosal and cutaneous barrier dysfunction, impaired antigen capture by dendritic cells, reduced cytotoxicity of natural killer (NK) cells, and diminished phagocytic activity of neutrophils [3]. Age-related effects on the adaptive immune system include a reduction in the proportion of CD3⁺ T cells, which decreases the total mature T-lymphocyte pool and impairs cell-mediated immunity. The decrease in the proportion of CD8⁺ cytotoxic T cells compromises viral clearance and tumor immunosurveillance, whereas shifts in the amount of CD4⁺ T-cell subsets impair helper functions, ultimately blunting antibody responses and vaccine efficacy [4, 5]. Another critical phenomenon in geriatric immunity is inflammaging, a state of chronic, low-grade systemic inflammation that develops during the aging process. It is characterized by elevated serum concentrations of C-reactive protein (CRP) and key proinflammatory cytokines, including interleukin (IL)-6, IL-8, and tumor necrosis factor-α (TNF-α). Studies have shown that inflammaging serves as a significant predictor of mortality risk among both healthy and frail elderly individuals. Conversely, lower levels of these proinflammatory markers are associated with greater longevity, better health outcomes, and a lower risk of death. Furthermore, inflammaging acts as a potent inhibitor of overall immune function and is recognized as one of the primary drivers of accelerated immunosenescence [6].

Probiotic supplementation represents a safe, noninvasive intervention to optimize the gut microbiota, bolster immunity, and reduce systemic inflammation [7,8]. In elderly individuals ( ≥ 65 years), probiotic supplementation enhances innate and adaptive defenses, thereby decreasing infection incidence and duration [9,10]. Specifically, probiotics bolster cellular immunity by increasing the count and function of NK cells, which act against viral and neoplastic cells that secrete critical cytokines such as IFN-γ [11]. Strains such as *Lactobacillus delbrueckii* subsp. *bulgaricus* OLL1073R-1 and *Bifidobacterium lactis* HN019 significantly increase NK cell cytotoxicity and lower the risk of the common cold in older adults [10,12]. Probiotics also expand T-cell subsets (e.g., CD3^+^, CD4^+^, and CD25^+^) and reduce upper respiratory tract infections, as observed for *B. lactis* HN019 and *Lactobacillus paracasei* N1115 [10,13]. To mitigate age-related chronic inflammation, strains such as *L. delbrueckii* subsp. *bulgaricus* 8481 and synbiotics (*Bifidobacterium longum* with Synergy 1™) significantly reduce the levels of proinflammatory cytokines such as IL-8 and TNF-α [14,15]. Finally, mucosal immune defenses are strengthened through an increased level of salivary secretory immunoglobulin A (sIgA) and salivary α-amylase (sAA) levels, both of which declines with age. Maintaining sIgA and sAA levels using probiotic strains like heat-killed *Lactobacillus pentosus* b240 demonstrates dose-dependent protection against cumulative cold incidence [9,16–19]. Thus, probiotic supplementation may represent a promising dietary strategy to mitigate age-related immune decline and protect against respiratory infections in older adults.

*Lactiplantibacillus plantarum* LP28 (*L. plantarum* LP28, formerly classified as *Lactobacillus plantarum*) was isolated from traditional Taiwanese dried tofu obtained from a traditional market in Daxi, Taoyuan City, Taiwan, and was supplied by SYNBIO TECH INC. (Kaohsiung, Taiwan). Preliminary observations from an unpublished internal study suggested the potential immunomodulatory effects of the LP28-containing formulation STP1 in a murine model of ovalbumin (OVA)-induced allergy. In this model, STP1 treatment was associated with changes in serum IgE levels and the secretion of IFN-γ and IL-10, providing preliminary evidence for its potential role in systemic immune modulation. Furthermore, an unpublished, small-scale exploratory study involving 48 adults receiving an LP28-containing formulation suggested potential benefits in terms of several common cold-related parameters. These included a lower frequency of recurrent colds, shorter durations of overall cold symptoms and persistent cough, and improvements in the perceived severity of the cold and associated symptoms such as headache and limb soreness. These observations provide preliminary support for further investigations of the potential effects of LP28 on upper respiratory tract health.

To address the growing health challenges associated with the global population aging, this study evaluated the effects of *L. plantarum* LP28 on the cold incidence and its immunomodulatory efficacy in middle-aged and elderly adults. Daily supplementation with *L. plantarum* LP28 markedly reduced both cold incidence and the cumulative symptom burden and strengthened innate immune defenses by elevating serum IL-12 levels and increasing NK cell cytotoxicity while simultaneously suppressing systemic inflammation through reduced CRP levels. Taken together, these findings demonstrate that *L. plantarum* LP28 supplementation is a viable nutritional strategy to bolster innate immunity, support healthy aging, and improve quality of life among middle-aged and elderly individuals.

## Materials and methods

### Recruitment of participants and study subjects

Middle-aged and older adults aged between 50 and 75 years were recruited. To ensure the study’s relevance to immune resilience, candidates needed to have experienced at least two episodes of the common cold or tonsillitis within the past year. Prior to enrollment, all potential participants were required to complete a comprehensive health screening questionnaire to evaluate their medical history. To minimize confounding variables and ensure participant safety, individuals meeting any of the following criteria were excluded from the study: those diagnosed by a physician with significant chronic or systemic diseases, including hepatic, renal, oncological, or autoimmune disorders; those with diagnosed psychiatric conditions, including but not limited to schizophrenia or clinical depression; individuals with chronic allergic rhinitis that could interfere with upper respiratory symptom assessments; those with a history of drug or alcohol dependency; and any individual deemed unsuitable for participation in a clinical trial by the lead investigator because of safety or compliance concerns. A total of 102 eligible participants provided informed consent and were enrolled in this randomized, double-blind, placebo-controlled trial.

### Study design

The participants were randomly assigned to either the probiotic group (*n* = 51) or the placebo group (*n* = 51) for a 12-week intervention period. Participants in the probiotic group consumed one probiotic capsule twice daily, within 30 minutes before breakfast and dinner. Each probiotic capsule contained 5 × 10⁹ colony-forming units (CFU) of *L. plantarum* LP28. Participants in the placebo group received capsules containing the same excipients (maltodextrin, microcrystalline cellulose, and magnesium stearate), without *L. plantarum* LP28, following the same dosing schedule. Prior to the intervention, all eligible participants underwent a 4-week washout period. During both the washout and 12-week intervention periods, participants were instructed to maintain their usual daily routines and dietary habits to minimize potential confounding from lifestyle and dietary changes. They were also instructed to strictly discontinue the use of immunosuppressants, steroids or glucocorticoids, respiratory health–targeted dietary supplements, probiotics, and fermented dairy products, including yogurt and probiotic drinks. Biological samples were collected at three time points: week 0 (baseline), week 6, and week 12 (endpoint). Fasting venous blood samples were collected for analysis of immune cell populations, NK cell cytotoxicity, cytokine levels, and clinical biochemistry and hematology parameters. Saliva samples were collected to measure amylase and sIgA levels. Throughout the 12-week trial, the participants were required to maintain daily and weekly health records, including completion of the Wisconsin Upper Respiratory Symptom Survey (WURSS-24), to monitor the incidence and severity of respiratory symptoms, as well as weekly documentation of vaccination status and common cold-related events. This study was conducted in accordance with the guidelines of the Declaration of Helsinki. The study protocol was reviewed and approved by the Institutional Review Board of Taichung Veterans General Hospital (IRB No. SF24108C). The clinical trial was registered at ClinicalTrials.gov under the registration number NCT06724185.

### Clinical Assessment and Patient-Reported Outcomes (PROs)

To systematically evaluate the participants’ health status and the efficacy of the intervention, three structured questionnaires were used throughout the trial. During the recruitment phase, a health screening questionnaire was administered to assess medical history and ensure that the candidates met the inclusion and exclusion criteria. During the study period, the participants completed daily questionnaires assessing upper respiratory symptoms. Upper respiratory symptoms were assessed using the Wisconsin Upper Respiratory Symptom Survey (WURSS), and summary scores were calculated to reflect the overall symptom burden. Cold occurrence was defined as the presence of at least one cold episode during the study period. Among participants who developed colds, daily discomfort was assessed using a self-reported discomfort score from the cold questionnaire. The number of discomfort days was defined as the number of days on which the discomfort score was greater than 0, reflecting the duration of subjective symptom burden. Symptom severity was further evaluated using the area under the curve (AUC) of discomfort scores over time, which was calculated using the trapezoidal rule and included only days with an observed discomfort score, the average discomfort score (AUC divided by the number of discomfort days) and the peak discomfort score (maximum daily discomfort score), which represented the cumulative burden, mean intensity during symptomatic periods, and highest symptom intensity, respectively. Simultaneously, the frequency and cumulative duration of common cold occurrences were documented. The logs included specific records of concurrent medication use during illness episodes and weekly updates on any vaccinations received, including the vaccine type and date of administration. These comprehensive patient-reported outcomes provided a longitudinal overview of the participants’ immune resilience in response to probiotic supplementation.

### Biochemical assessment and safety monitoring

At baseline (week 0) and at the study endpoint (week 12), all participants underwent physical examinations that included assessments of height, weight, and blood pressure. Additionally, 8 mL of the 38 mL venous blood sample collected from each participant was set aside for routine laboratory testing. Safety assessments included a complete blood count (CBC), including assessments of white blood cell (WBC) and red blood cell (RBC) counts, hemoglobin (Hb) levels, hematocrit (Hct), platelet (PLT) counts, and red cell indices (MCV, MCH, MCHC, RDW-SD), as well as liver function tests (measurement of AST, ALT, and γ-GT levels) and renal function tests (measurement of BUN, creatinine, eGFR, and uric acid levels). The clinical parameters quantified by these assessments served as critical safety indicators to monitor for any potential adverse reactions associated with the daily consumption of either the probiotic or placebo capsules throughout the trial period.

### Isolation and quantification of peripheral blood mononuclear cells (PBMCs)

Given the relatively low percentage of NK cells among PBMCs, a starting volume of 30 mL of fresh venous blood was collected to ensure sufficient cell yield for downstream functional assays. The initial step of the isolation process was centrifugation at room temperature for 10 minutes to separate the plasma. The resulting supernatant was carefully aliquoted into microcentrifuge tubes and stored at -80 °C for subsequent systemic cytokine profiling. The remaining blood fraction was then diluted with an equal volume (approximately 12 to 15 mL) of sterile phosphate-buffered saline (PBS; Gibco, Carlsbad, CA, USA). To isolate the mononuclear fraction, the diluted blood was gently layered over 15 mL of Ficoll®-Paque Plus (Sigma, St. Louis, MO, USA) density gradient medium in a centrifuge tube, taking care to maintain a sharp interface between the layers. Following centrifugation at room temperature for 30 minutes, the PBMCs were meticulously aspirated using a sterile pipette and transferred to a new conical tube. The collected PBMCs were then washed and resuspended in an equal volume of PBS. Finally, the cell concentration and cell viability were determined using the trypan blue exclusion method; specifically, 20 μL of the cell suspension was mixed with an equal volume of 0.4% trypan blue solution and loaded onto a hemocytometer for manual counting. This procedure ensured a high-purity cell population with verified viability for subsequent immunophenotyping and NK cell purification.

### Immune cell analysis

To characterize the lymphocyte subpopulations, 2 × 10^5^ isolated PBMCs were allocated for each immunophenotyping panel. The cell suspension was first centrifuged, and the supernatant was discarded, after which extracellular staining was performed. Specifically, fluorochrome-conjugated monoclonal antibodies, including anti-CD3-APC, anti-CD4-PE, anti-CD8-FITC, anti-CD16-APC, and anti-CD56-FITC (BioLegend, San Diego, CA, USA), were diluted 1:100 in staining buffer consisting of PBS supplemented with 1% fetal bovine serum (FBS; Gibco). The cells were incubated with an antibody cocktail on ice for one hour in the dark to facilitate optimal binding while minimizing fluorophore degradation. After incubation, the cells were washed with 1 mL of PBS and centrifuged to remove the unbound antibodies. The resulting cell pellets were resuspended in 600 μL of PBS for data acquisition, which was performed using a BD FACSCanto II flow cytometer (BD Biosciences, San Jose, CA, USA). During analysis, lymphocytes were initially gated based on their characteristic light-scattering properties, specifically forward scatter and side scatter, which reflect cell size and granularity, respectively. The fluorescence signals emitted from specific cell-surface markers were subsequently analyzed to quantify the proportions of T cells and NK cells within the total gated population of lymphocytes.

### Purification of NK cells and cytotoxicity assay

NK cells were purified from the isolated PBMCs using a magnetic-bead-based negative selection isolation kit (STEMCELL Technologies, Cambridge, MA, USA). This method ensured the enrichment of untouched NK cells through the depletion of nontargeting cells. The PBMCs were transferred to a 5 mL round-bottom sterile tube, to which 50 μL of the isolation antibody cocktail was added, and the mixture was incubated at room temperature for 5 minutes. Afterward, 50 μL of RapidSpheres™ magnetic particles were added, followed by the immediate addition of 1.5 mL of PBS. The mixture was then placed in a magnetic separator for 3 minutes. During this period, magnetically labeled non-NK cells were sequestered against the tube wall, allowing the purified NK cell suspension to be decanted into a centrifuge tube containing 10 mL of RPMI-1640 medium (Gibco). The enriched NK cells were harvested via centrifugation at room temperature for 5 minutes, resuspended in 1 mL of fresh RPMI medium, and quantified using a hemocytometer, with the trypan blue exclusion method used to verify total cell yield and viability. K562 human immortalized myelogenous leukemia cells (CCL-243, ATCC, Manassas, VA, USA) served as the target cells. These target cells (1×10^4^ cells/well) were cocultured with effector NK cells at specific effector-to-target (E:T) ratios of 1:1 (1 × 10^4^ NK cells) and 5:1 (5 × 10^4^ NK cells) for 4 hours. Following incubation, the cell culture supernatant was harvested and analyzed using a nonradioactive lactate dehydrogenase (LDH) detection kit (Abcam, Cambridge, UK). The concentration of LDH, an enzyme released into the medium upon plasma membrane damage, was measured colorimetrically. This quantitative value represented the proportion of K562 cells that had been lysed by NK cells and served as a definitive indicator of the cytotoxic activity of NK cells.

### Systemic cytokine concentration and salivary immune marker quantification

To quantify systemic immune responses, inflammatory status, and mucosal immunity, plasma and saliva samples were collected at baseline (week 0) and at weeks 6 and 12. The concentrations of cytokines, including IL-4, IL-6, IL-12, IFN-γ, CRP, and TNF-α, were analyzed in the plasma samples, whereas sIgA and α-amylase levels were measured in the salivary samples. The plasma and saliva samples were stored at −80 °C until analysis. Prior to testing, the plasma samples were thawed on ice, whereas the saliva samples were thawed on ice and centrifuged at 4 °C for 10 minutes to remove mucins and cellular debris, yielding clarified supernatants. The levels of all analytes were quantified using high-sensitivity human enzyme-linked immunosorbent assay (ELISA) kits (Elabscience, Houston, TX, USA) according to the manufacturer’s instructions. Briefly, clarified samples (100 μL per well) were loaded in triplicate into precoated microplates, along with serially diluted standards, to generate standard curves. The plates were incubated at 4 °C overnight to allow antigen–antibody binding, after which the unbound materials were removed. Biotinylated detection antibodies and enzyme-conjugated secondary antibodies (streptavidin-HRP) were sequentially added, and the samples were incubated at room temperature. After the final wash, a chromogenic substrate (TMB) was added, and the samples were incubated in the dark for 15 minutes to allow color development. The reaction was terminated using a stop solution, and the optical density (OD) was measured at 450 nm using a microplate reader (Thermo Fisher Scientific, Boston, MA, USA). The concentrations of cytokines and salivary proteins were calculated using linear regression equations derived from the corresponding standard curves and are expressed as pg/mL or ng/mL.

### Statistical analysis

Continuous variables are presented as mean ± standard deviation and were compared using independent *t* tests or paired *t* tests, as appropriate. Categorical variables were analyzed using chi-square tests or Fisher’s exact tests. Data interpretation was performed following two distinct strategies: outlier-removed sensitivity analysis and a refined per-protocol analysis to consider physiological variability and extreme outliers. The per-protocol (PP) population included participants who completed the 12-week intervention without major protocol deviations. To evaluate the robustness of the biological marker data (e.g., serum cytokine concentrations), an outlier-removed sensitivity analysis was performed by excluding extreme values (those exceeding 2.5 standard deviations from the group mean). For analyses restricted to participants who developed colds, between-group comparisons were performed using the Mann–Whitney *U* test because of the small sample size and nonnormal data distribution. Odds ratios (ORs) with 95% confidence intervals (CIs) are reported to evaluate symptom dynamics (probiotic vs. placebo, with an OR < 1 indicating a faster decline in symptom incidence in the probiotic group). Multiple comparisons across symptoms were adjusted using the false discovery rate (FDR) method. A two-sided *P* value < 0.05 was considered to indicate statistical significance.

## Research results

### Participant flow and sample representativeness

A total of 102 volunteers were initially recruited for this study (**Figure 1**) and randomly assigned to the placebo group (*n* = 51) or the probiotic group (*n* = 51). To ensure the integrity and accuracy of the longitudinal data, participant status was closely monitored throughout the trial. During the intervention, six participants were excluded: one in the placebo group (due to illness) and five in the probiotic group (one due to illness, two due to refusal to undergo a second blood test, and two due to missing questionnaire data). Consequently, 96 participants successfully completed all clinical assessments from baseline to the study endpoint, with 50 participants in the placebo group (*n* = 50) and 46 participants in the probiotic group (*n* = 46). Although the final sample size (*n* = 46 and *n* = 50) provided 80% power to detect moderate-to-large between-group differences (Cohen’s d ≥ 0.58; two-sided α = 0.05), it was underpowered for the observed difference in cold occurrence (estimated power ≈ 40%).

**Figure 1.**
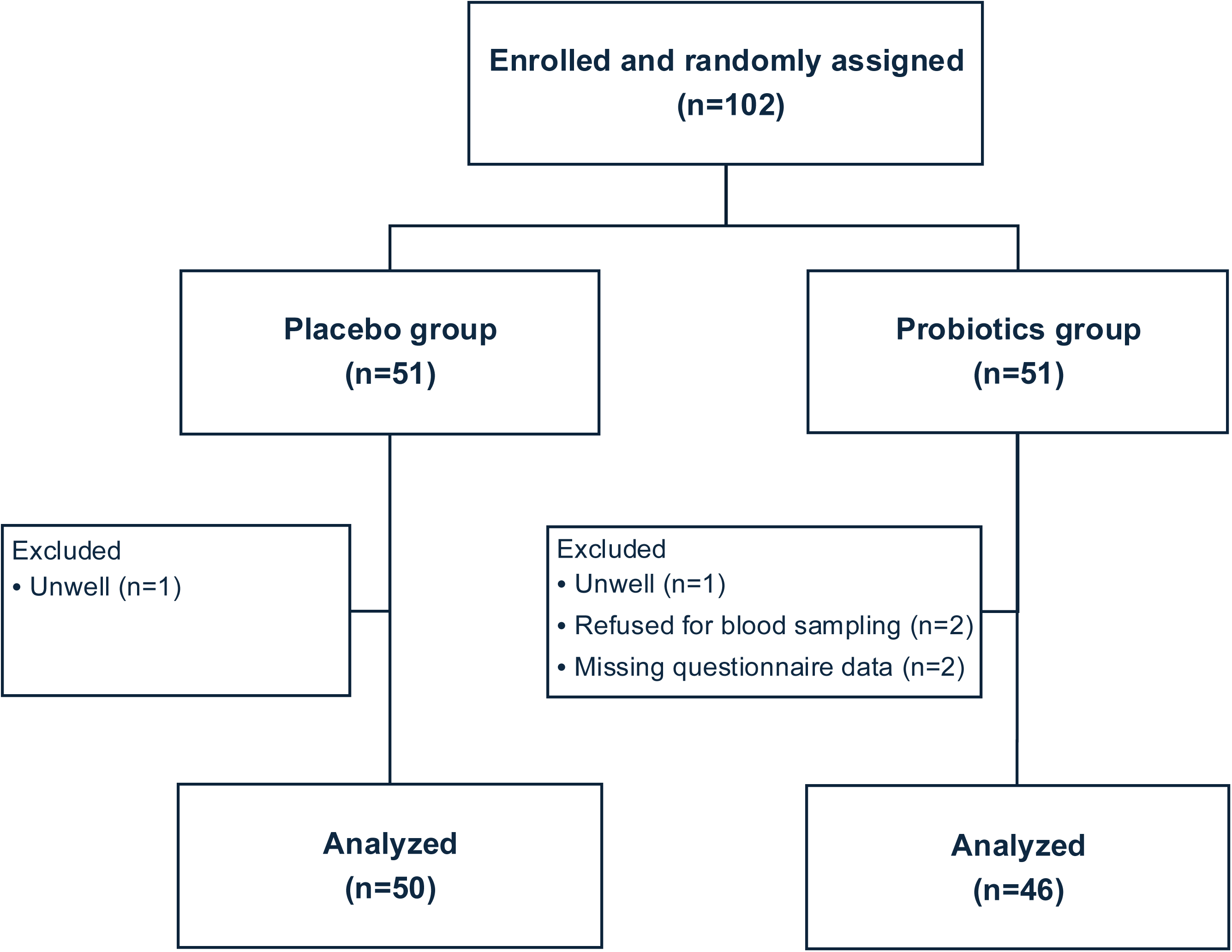
Flowchart of study participant selection. The final analysis was performed following a per-protocol (PP) approach, excluding participants with incomplete data or extreme physiological outliers to ensure that the biological efficacy of the intervention was accurately captured.

The mean age of the participants was 62.67 ± 8.82 years in the probiotic group and 60.72 ± 7.89 years in the placebo group. Height and body weight were also comparable between the groups. The distributions of the continuous variables were approximately normal, as indicated by the skewness and kurtosis values. There were no significant differences between the probiotic and placebo groups in terms of baseline characteristics, including age (*p* = 0.255), height (*p* = 0.455), or body weight (*p* = 0.502). All effect sizes were small (Cohen’s d < 0.25), indicating negligible between-group differences (**Table 1**).

**Table 1.**
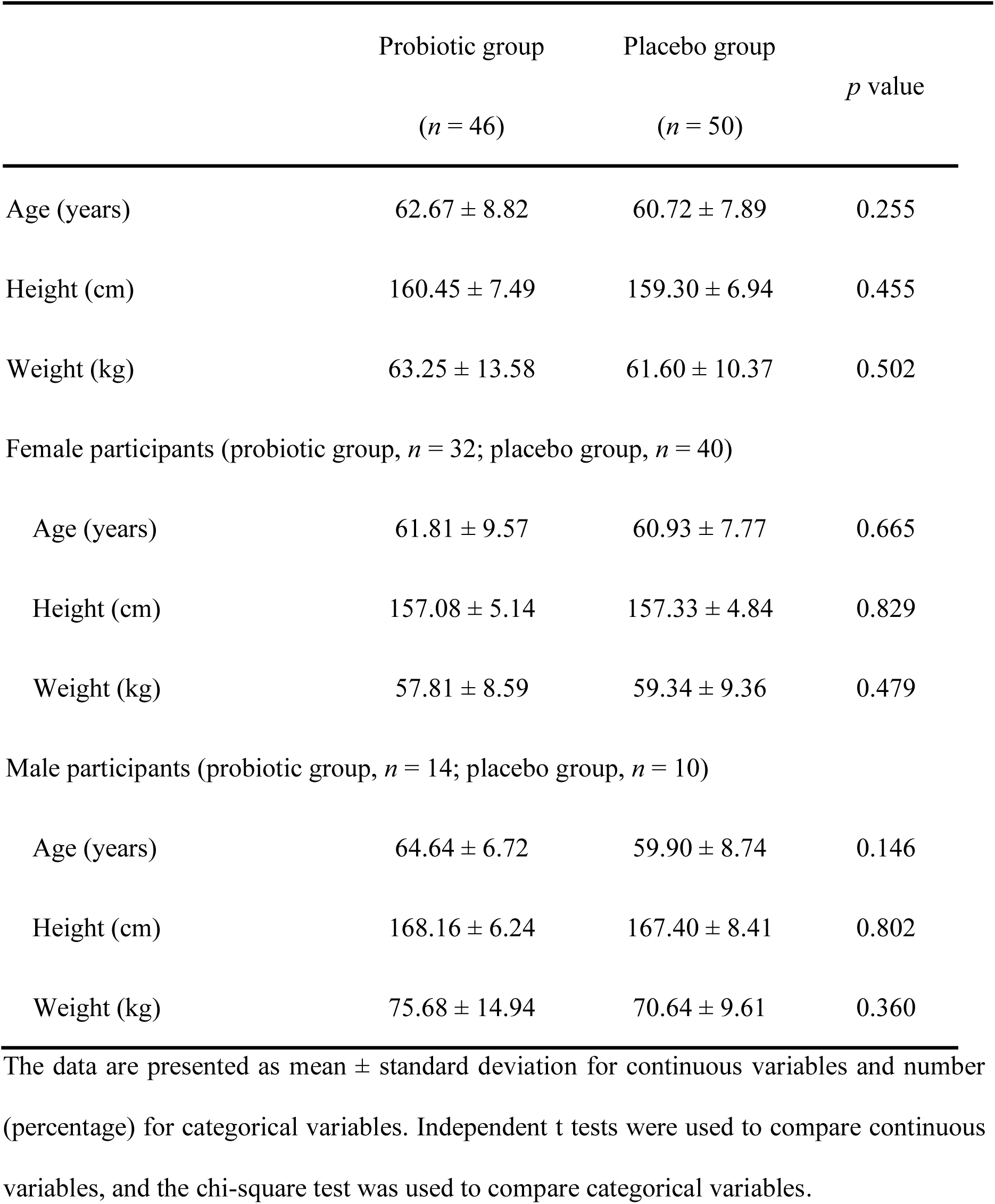
Baseline characteristics of participants.

### Potential of *L. plantarum* LP28 supplementation in reducing cold incidence and modulating upper respiratory symptoms

During the 12-week study period, the proportion of participants who experienced at least one cold episode was lower in the probiotic group than in the placebo group (34.78% [16/46] vs. 52.00% [26/50]; OR = 2.02; 95% CI: 0.83–5.03; Fisher’s exact test, *p* = 0.103), suggesting a potential protective effect of the intervention; however, the difference did not reach statistical significance (**Table 2**). Among participants who developed colds (infection-only analysis), no statistically significant differences were observed between the probiotic and placebo groups in terms of the number of discomfort days, average discomfort score, peak discomfort score, or AUC (all *p* > 0.20; **Table 2**). The mean number of discomfort days was 8.50 ± 11.34 in the probiotic group and 13.62 ± 23.15 in the placebo group (*p* = 0.397). The mean discomfort score was 8.58 ± 9.47 in the probiotic group vs. 9.30 ± 7.26 in the placebo group (*p* = 0.524), the peak discomfort score was 14.67 ± 16.39 vs. 25.48 ± 32.67 (*p* = 0.320), and the AUC was 72.00 ± 127.37 vs. 98.48 ± 129.87 (*p* = 0.231), respectively. Although these differences did not reach statistical significance, the effect size estimates (Cliff’s δ ranging from −0.14 to −0.26) consistently favored the probiotic group, suggesting a small tendency toward lower symptom burden among participants receiving probiotic supplementation (**Table 2**).

**Table 2.** Effects of *L. plantarum* LP28 supplementation on cold occurrence and symptom severity.

|  | Probiotic group | Placebo group | *p* value | Effect size (Cliff’s δ) |
|---|---|---|---|---|
| Cold occurrence (%) | 34.78% (16/46) | 52.00% (26/50) | 0.103 | OR = 2.02 (95% CI: 0.83-5.03) |
| Among participants with cold (infection-only analysis) |  |  |  |  |
| Number of discomfort days | 8.50 ± 11.34 | 13.62 ± 23.15 | 0.397 | -0.18 |
| Discomfort score (AVG) | 8.58 ± 9.47 | 9.30 ± 7.26 | 0.524 | -0.14 |
| Discomfort score (peak) | 14.67 ± 16.39 | 25.48± 32.67 | 0.320 | -0.21 |
| Discomfort score (AUC) | 72.00 ± 127.37 | 98.48 ± 129.87 | 0.231 | -0.26 |

The data are presented as mean ± standard deviation for symptom-related outcomes. Cold occurrence was calculated using the full cohort, whereas symptom-related outcomes were analyzed only among participants who developed at least one cold episode during follow-up. Between-group comparisons for symptom-related outcomes were performed using the Mann–Whitney U test. Effect sizes are presented as Cliff’s delta; negative values indicate lower values in the probiotic group. Odds ratios (ORs) are presented for the probiotic group versus the placebo group. Symptom-related analyses were conducted on an available-case basis; observations for which values for a given outcome were missing were excluded from the corresponding analysis, and missing values were not imputed.

The heatmap demonstrated that cumulative WURSS-24 symptom scores were generally lower in the probiotic group across multiple symptom domains (**Figure 2A**). The forest plot revealed odds ratios less than 1.0 for most symptoms, suggesting a possible protective effect (**Figure 2B**). However, the confidence intervals were wide and frequently included 1.0; thus, the results did not provide clear evidence of reduced odds for these symptoms.

**Figure 2.**
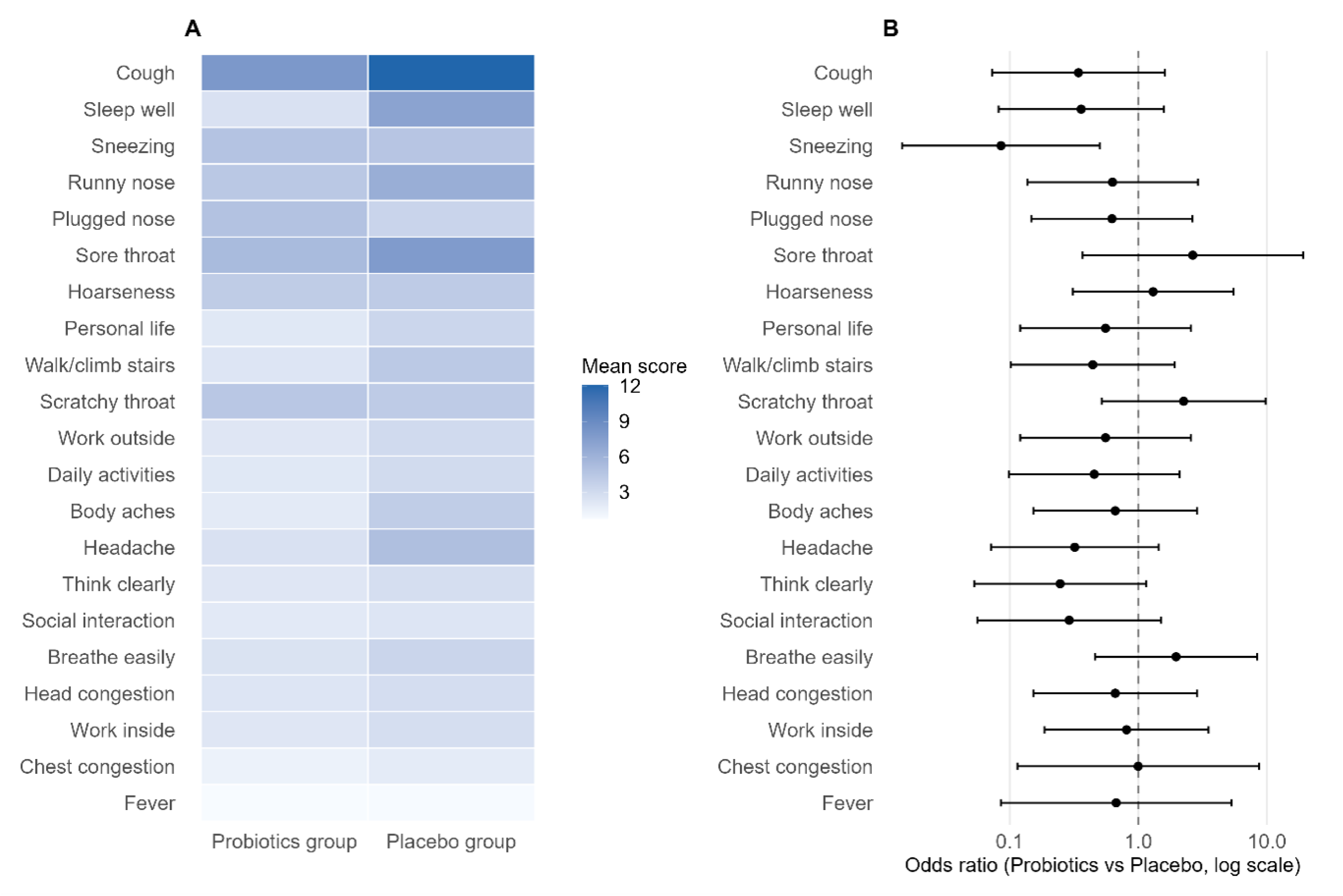
Comparison of symptom severity scores and odds ratios for symptom incidence between the *L. plantarum* LP28 (probiotic) and placebo groups. (A) Heatmap illustrating mean cumulative symptom severity according to the WURSS and functional impact scores. Darker shades of blue represent greater severity or greater functional impairment. (B) Forest plot showing the odds ratios (ORs) with 95% confidence intervals for symptom incidence (probiotic group vs. placebo group). An OR < 1.0 indicated a protective effect of the intervention.

### *L. plantarum* LP28 supplementation maintains immune cell homeostasis

To evaluate whether long-term probiotic supplementation modulates the overall composition of the systemic immune cell population, we assessed the immune cell population using flow cytometry. In the placebo group, a notable trend was observed: the proportion of CD3^+^ T cells was significantly increased after 12 weeks of intervention compared with that at baseline (*p* = 0.0428) (**Figure 3A**). Concurrently, the proportion of CD3^+^CD4^+^ T cells (helper T cells) increased, but the difference was not statistically significant (*p* = 0.0713) (**Figure 3B**). In contrast, the percentages of CD3^+^CD8^+^ T cells (cytotoxic T cells) and CD16^+^CD56^+^ NK cells remained stable in the placebo group (**Figure 3C and 3D**). Conversely, the immunological profile of the probiotic group was highly stable. Statistical analysis revealed no significant difference in the proportion of CD3^+^ T cells, CD3^+^CD4^+^ T cells, CD3^+^CD8^+^ T cells, or CD16^+^CD56^+^ NK cells between the 6-week or 12-week time point and baseline (**Figure 3A–D**). These observations were further confirmed by a percentage change analysis, which revealed that the proportions of CD3^+^ T cells, CD3^+^CD4^+^ T cells, CD3^+^CD8^+^ T cells, and CD16^+^CD56^+^ NK cells remained near baseline levels at all time points in both groups (**Figures 3E–H**). These findings suggested that the specific probiotic strain utilized in this study did not disrupt the homeostatic distribution of major immune cell subsets in healthy individuals, thereby supporting the maintenance of internal physiological stability.

**Figure 3.**
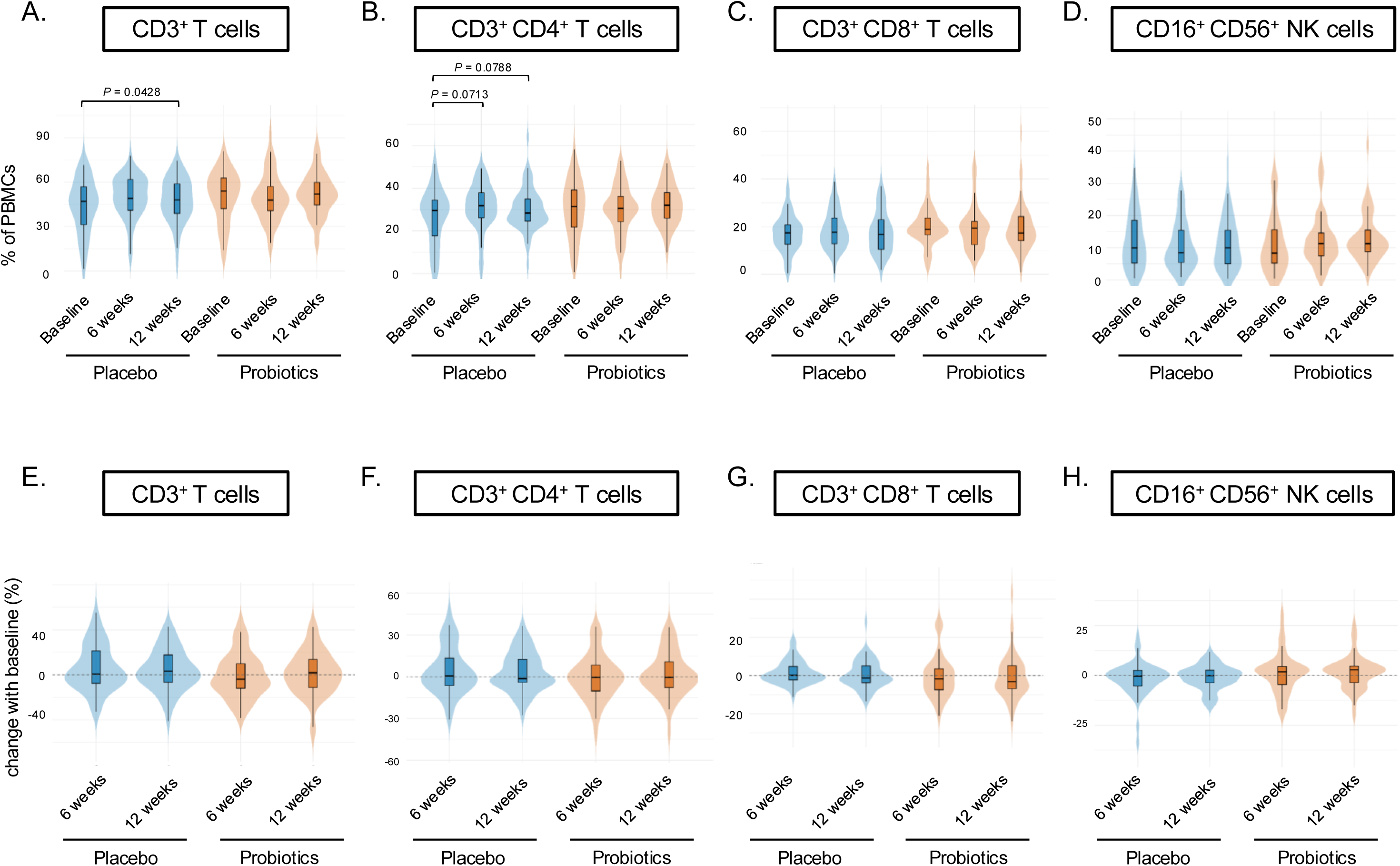
Effects of *L. plantarum* LP28 supplementation on the composition of systemic immune cell populations. The proportions of major immune cell subsets were analyzed using flow cytometry at baseline, week 6, and week 12. (**A**) Percentage of total CD3^+^ T cells. (**B**) Percentage of CD3^+^CD4^+^ helper T cells. (**C**) Percentage of CD3^+^CD8^+^ cytotoxic T cells and (**D**) percentage of CD16^+^CD56^+^ NK cells. The distribution of percentage changes from baseline for (**E**) CD3^+^ T cells, (**F**) CD3^+^CD4^+^ T cells, (**G**) CD3^+^CD8^+^ T cells, and (**H**) CD16^+^CD56^+^ NK cells. These violin plots represent the individual variability and the stability of change relative to the baseline (indicated by the dashed line at 0%). The data are presented as mean ± SD.

### *L. plantarum* LP28 supplementation increases NK cell cytotoxic activity

Although the relative proportions of immune cell subsets remained largely unchanged (**Figure 3**), the experimental data further confirmed that probiotic supplementation exerted a significant modulatory effect on the functional activity of immune cells. As NK cells are the primary line of defense in the innate immune system, their cytotoxic activity serves as a critical indicator for assessing overall immune competence. Our results demonstrated that participants in the probiotic group exhibited a significant and sustained increase in NK cell cytotoxicity at both 6 and 12 weeks of intervention compared baseline when the E/T ratio was 1:1 (baseline vs. 6 weeks, *p* = 0.0058; baseline vs. 12 weeks, *p* = 0.006) (**Figure 4A**). Consistent with these findings, when the E/T ratio was 5:1, NK cell cytotoxicity was significantly higher at weeks 6 and 12 than at baseline (baseline vs. 6 weeks, *p* = 0.005; baseline vs. 12 weeks, *p* = 0.0009) (**Figure 4C**). More importantly, intergroup analysis revealed that compared with that in the placebo group, NK cell activity in the probiotic group was significantly greater at the 6-week time point (E/T =1, placebo vs. probiotic, *p* =0.0232; E/T =5, placebo vs. probiotic, *p* =0.0203), but there was no significant difference at the 12-week time point for either the 1:1 or 5:1 E/T ratio (E/T =1, placebo vs. probiotic, *p* =0.0623; E/T =5, placebo vs. probiotic, *p* =0.4139) (**Figure 4B and 4D**). Collectively, these data suggested that the probiotic exerted beneficial immunomodulatory effects primarily by increasing the functional efficiency of NK cells rather than altering their systemic distribution.

**Figure 4.**
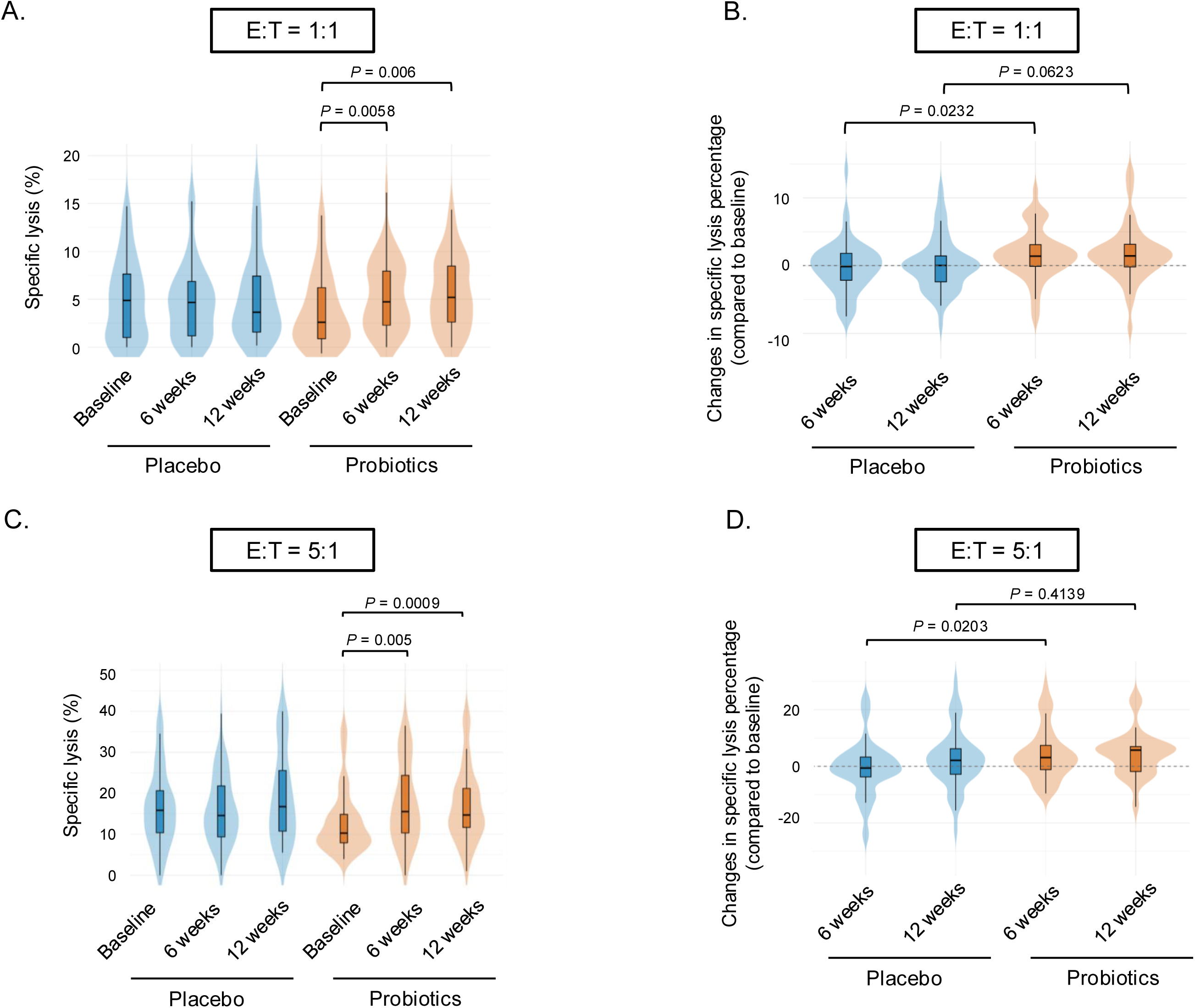
Enhancement of NK cell cytotoxicity following *L. plantarum* LP28 supplementation. NK cell activity was evaluated by measuring cytotoxic potential against target cells at baseline, week 6, and week 12. **(A)** Intragroup changes in NK cell cytotoxicity at an effector-to-target (E/T) ratio of 1:1 across time points. **(B)** Intergroup comparison of NK cell cytotoxicity between the probiotic and placebo groups at an E/T ratio of 1:1. **(C)** Intragroup changes in NK cell cytotoxicity at an E/T ratio of 5:1. **(D)** Intergroup comparison of NK cell cytotoxicity between the probiotic and placebo groups at an E/T ratio of 5:1. The data are presented as mean ± SD.

### Modulation of selective cytokine concentrations and immune markers by *L. plantarum*

#### LP28 supplementation

To further investigate the systemic and mucosal immune modulatory effects of probiotic supplementation, we quantified the levels of key cytokines and proteins in both serum and saliva. In the probiotic group, IL-12 levels were significantly increased at the 6-week time point compared to baseline (baseline vs. 6 weeks, *p* = 0.0366) (**Figure 5C**). Concurrently, a significant reduction in CRP levels was observed at week 6, suggesting a potential anti-inflammatory effect or a shift in systemic inflammation (baseline vs. 6 weeks, *p* = 0.024) (**Figure 5F**). However, there was no significant difference in the change from baseline in the IL-12 (baseline vs. 12 weeks, *p* = 0.1071) or CRP (baseline vs. 12 weeks, *p* = 0.1502) level between the probiotic and placebo groups during the 12-week intervention period (**Figure 5K** and **5N**). With respect to serum IFN-γ levels, no statistically significant differences were observed in the probiotic group at week 6 or week 12 compared with baseline, although an increasing trend was observed at week 12 (**Figure 5E and 5M**). The levels of other markers, including serum IL-4 (**Figure 5A and 5I**), IL-6 (**Figure 5B and 5J**), and TNF-α (**Figure 5D and 5L**), as well as the salivary sIgA level (**Figure 5H and 5P**), remained similar to those at baseline throughout the 12-week period. In contrast, the placebo group exhibited notable stability across most markers, including serum IFN-γ levels (**Figure 5E**), with the only exception being the salivary α-amylase level, which was significantly lower at week 6 than at baseline (*p* = 0.034) (**Figure 5G**). However, this decrease was transient, as the concentrations returned to baseline levels by week 12. No other statistically significant changes were observed in either serum cytokine or salivary sIgA levels within the placebo group. Taken together, these results suggested that probiotics may function as immunomodulators, effectively ’priming’ the IL-12/NK cell axis while mitigating systemic inflammation. Crucially, the maintenance of the levels of other cytokines confirmed that this functional enhancement was achieved without inducing unnecessary inflammatory stress or physiological imbalance.

**Figure 5.**
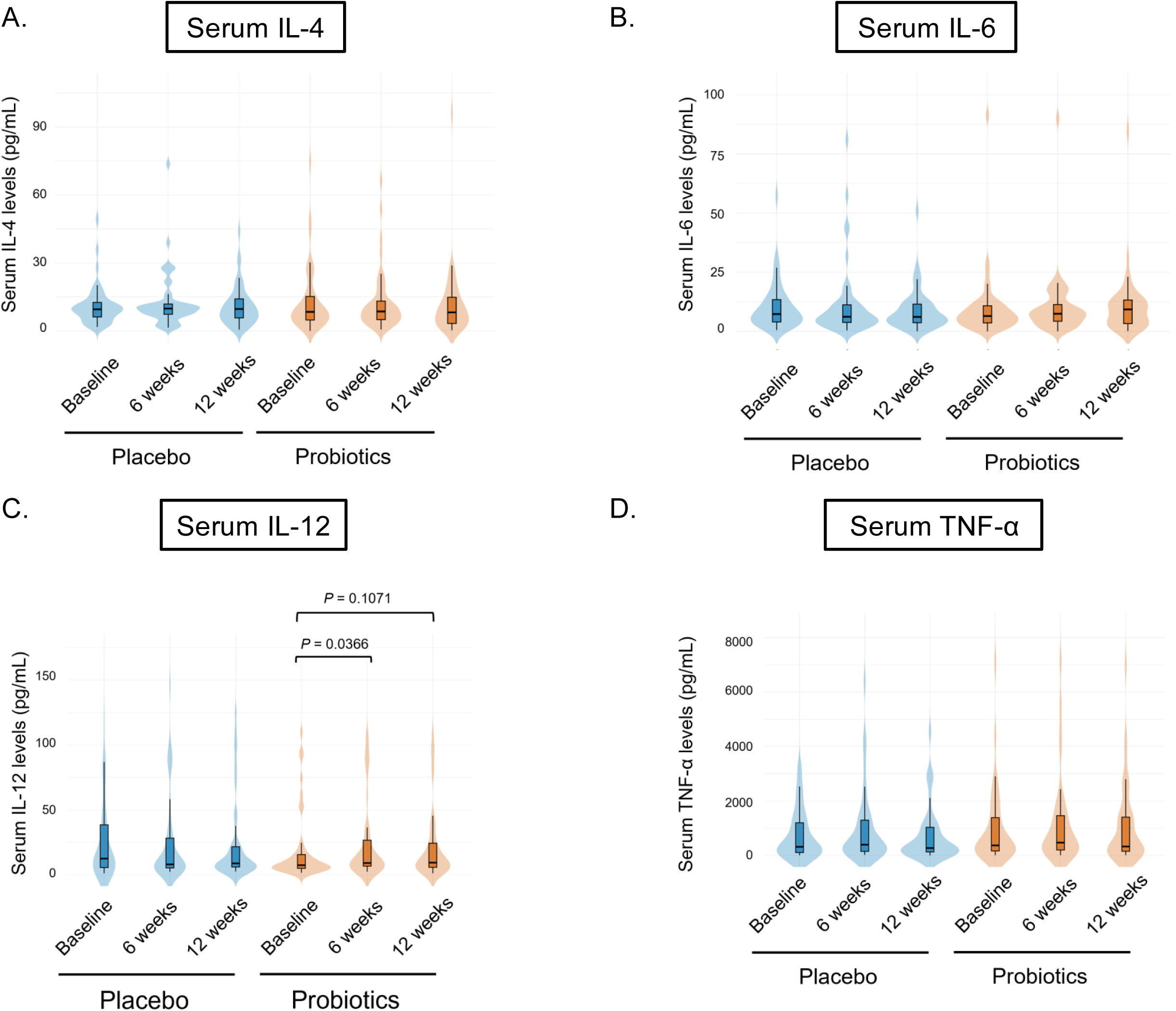
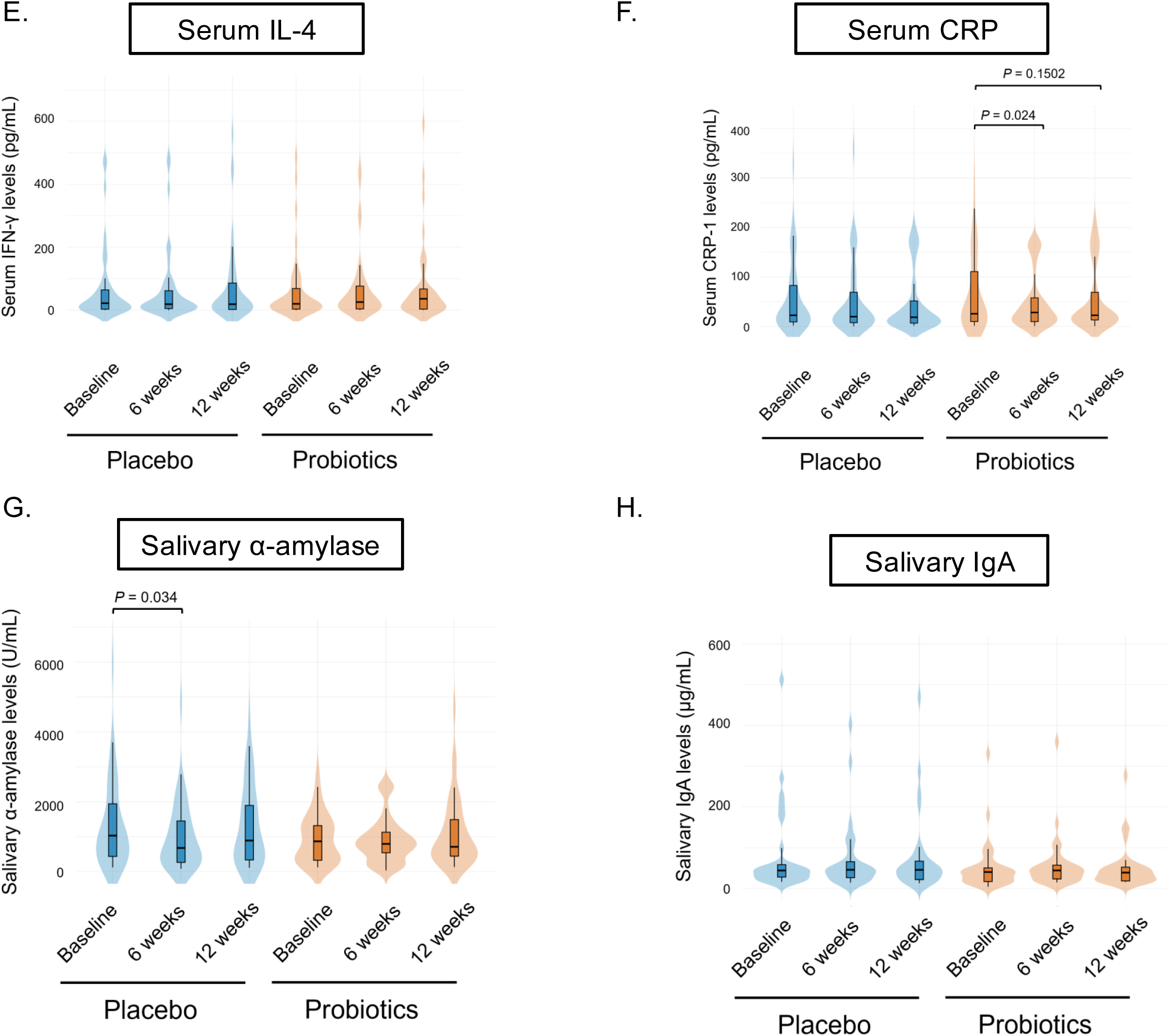
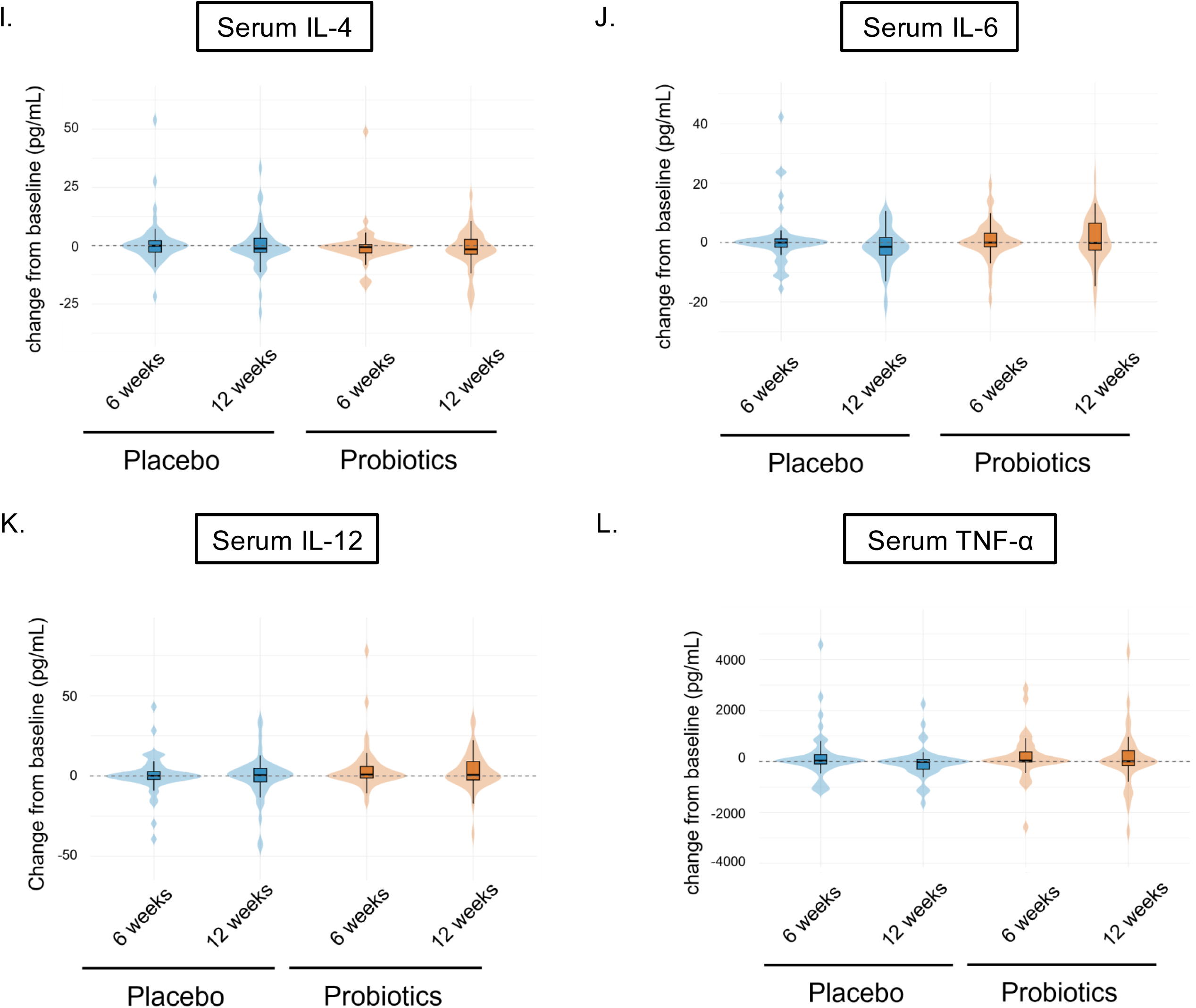
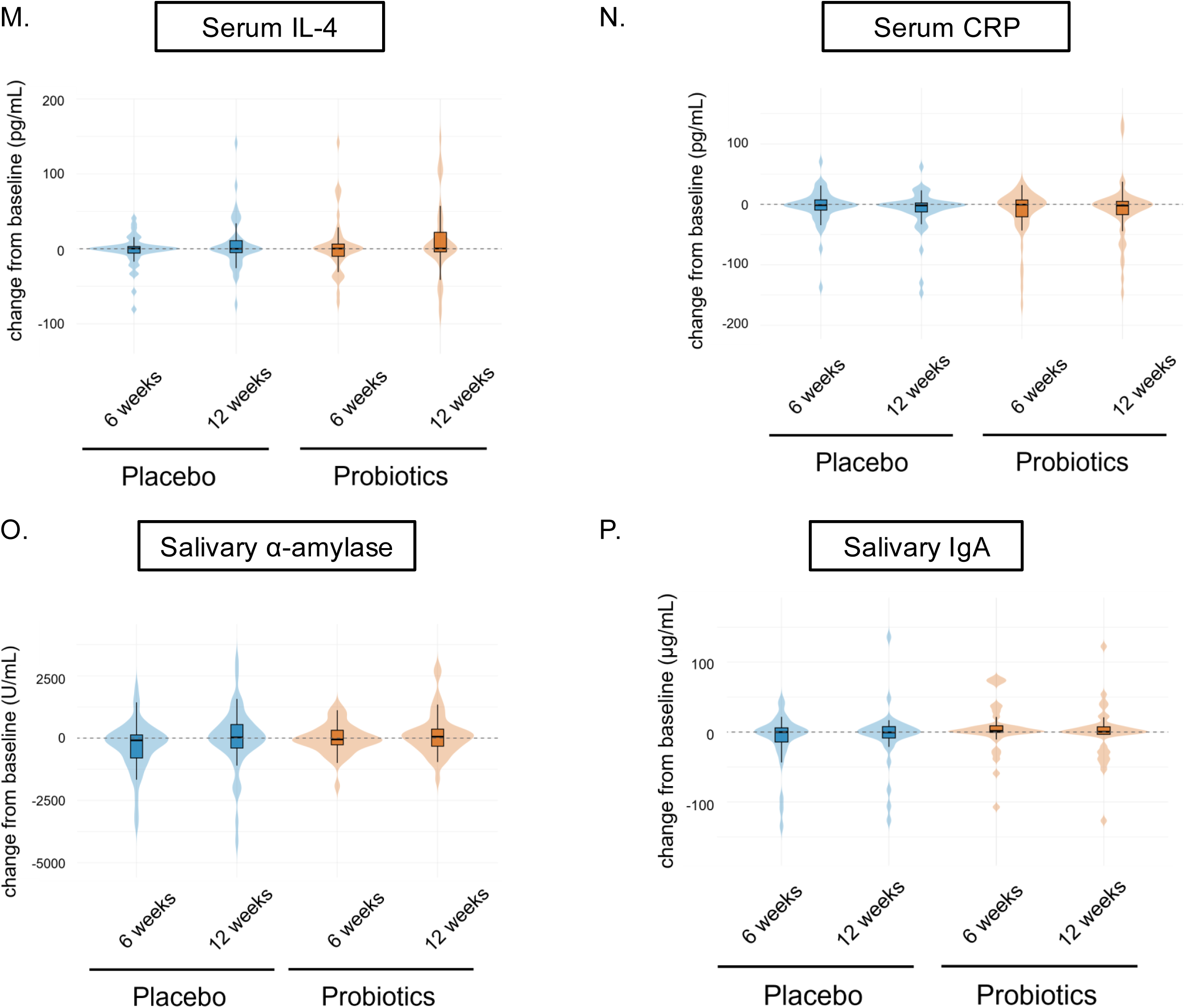
Modulation of selective cytokine levels and immune markers in serum and saliva by *L. plantarum* LP28 supplementation. (A–F) Longitudinal profiles of serum levels of cytokines and inflammatory markers, including IL-4, IL-6, IL-12, TNF-α, IFN-γ, and CRP, measured at baseline, week 6, and week 12. **(G, H)** Salivary mucosal levels of immune markers, specifically α-amylase and secretory IgA (sIgA), across time points. **(I–P)** Violin plots depicting the distribution of percentage changes from baseline (%) for the levels of serum cytokines (IL-4, IL-6, IL-12, TNF-α, IFN-γ, and CRP) and salivary markers (α-amylase and sIgA) at weeks 6 and 12. The dashed lines in the violin plots represent baseline parity (0% change). The data are presented as mean ± SD. Significant differences (\**p* < 0.05) are relative to the respective baseline values.

#### Safety and tolerability of *L. plantarum* LP28 supplementation

Finally, to ensure the clinical safety and tolerability of long-term probiotic supplementation, a comprehensive panel of hematological and biochemical markers was monitored (**Table 3**). Throughout the 12-week study period, all measured parameters, including hepatic and renal function indicators, remained within established normal physiological ranges for all participants. Although a statistically significant decrease in WBC count was noted in the probiotic group and significant fluctuations in the amount of MCH were observed in both groups, these variations remained well within healthy clinical reference limits and were not considered clinically adverse. No significant disturbances in the levels of hepatic enzymes (AST, ALT, or γ-GT) or renal function markers (BUN, creatinine, or eGFR) were detected. Collectively, these findings demonstrated that daily intake of *L. plantarum* LP28 was safe and maintained systemic physiological homeostasis without inducing metabolic or hematological toxicity.

**Table 3.** Safety assessment using hematological and clinical biochemical parameters during the 12-week intervention.

| Parameters | Group | Baseline<br>(Mean ± SD) | Week 12<br>(Mean ± SD) | *p* value<br>(intragroup) |
|---|---|---|---|---|
| **Hematology** |  |  |  |  |
| WBC (1 x 10<sup>3</sup> cells/ μ L) | Placebo | 5.5 ± 1.3 | 5.4 ± 1.3 | 0.8212 |
|  | Probiotic | 5.8 ± 1.7 | 5.3 ± 1.5 | 0.0379* |
| RBC (1 x 10<sup>6</sup> cells/μL) | Placebo | 4.6 ± 0.6 | 4.6 ± 0.6 | 0.5625 |
|  | Probiotic | 4.5 ± 0.4 | 4.6 ± 0.4 | 0.0714 |
| HGB (g/dL) | Placebo | 13.3 ± 1.1 | 13.4 ± 1.0 | 0.4038 |
|  | Probiotic | 13.7 ± 1.2 | 13.7 ± 1.0 | 0.6192 |
| HCT (%) | Placebo | 39.8 ± 3.1 | 40 ± 3.0 | 0.5413 |
|  | Probiotic | 40.5 ± 3.5 | 40.7 ± 2.8 | 0.552 |
| MCV (fL) | Placebo | 87.7 ± 7.7 | 87.7 ± 8.0 | 0.9294 |
|  | Probiotic | 89.2 ± 3.3 | 88.5 ± 4.2 | 0.0732 |
| MCH (pg) | Placebo | 31.0 ± 3.1 | 30 ± 3.3 | 0.0117* |
|  | Probiotic | 31.8 ± 2.0 | 30.7 ± 2.2 | 0.0087* |
| MCHC (g/dL) | Placebo | 33.5 ± 0.8 | 33.6 ± 0.8 | 0.7346 |
|  | Probiotic | 33.7 ± 0.6 | 33.7 ± 0.7 | 0.7759 |
| Platelet (1 x 10<sup>3</sup> cells/μL) | Placebo | 253.7 ± 47.6 | 250.7 ± 58.0 | 0.5872 |
|  | Probiotic | 256.8 ± 98.0 | 247.7 ± 91.1 | 0.1435 |
| RDW-CV (%) | Placebo | 13.8 ± 0.9 | 13.7 ± 1.0 | 0.3971 |
|  | Probiotic | 13.3 ± 0.5 | 13.4 ± 0.7 | 0.5281 |
| **Liver Function Tests** |  |  |  |  |
| AST (U/L) | Placebo | 22.9 ± 9.9 | 22.1 ± 6.6 | 0.255 |
|  | Probiotic | 23.2 ± 6.8 | 22.5 ± 6.0 | 0.2647 |
| ALT (U/L) | Placebo | 23.9 ± 21.6 | 22.2 ± 16.7 | 0.1503 |
|  | Probiotic | 21.2 ± 10.2 | 20.8 ± 9.0 | 0.6769 |
| r-GT (U/L) | Placebo | 18.0 ± 10.3 | 16.9 ± 11.8 | 0.3862 |
|  | Probiotic | 20.6 ± 12.5 | 19.5 ± 9.2 | 0.3095 |
| **Renal Function Tests** |  |  |  |  |
| BUN (mg/dL) | Placebo | 14.6 ± 3.3 | 13.9 ± 2.9 | 0.0984 |
|  | Probiotic | 14.8 ± 3.7 | 15.4 ± 5.3 | 0.3195 |
| Creatinine (mg/dL) | Placebo | 0.8 ± 0.2 | 0.8 ± 0.2 | 0.7761 |
|  | Probiotic | 0.8 ± 0.2 | 0.8 ± 0.2 | 0.296 |
| MDRD-based eGFR<br>(mL/min/1.73m<sup>2</sup>) | Placebo | 92.8 ± 20.5 | 90.2 ± 16.2 | 0.175 |
|  | Probiotic | 85.5 ± 16.7 | 88.1 ± 21.0 | 0.1665 |
| Uric acid (mg/dL) | Placebo | 5.0 ± 1.2 | 4.9 ± 1.3 | 0.3133 |
|  | Probiotic | 5.3 ± 1.3 | 5.2 ± 1.3 | 0.1799 |

The data are presented as mean ± SD. *p* values (intragroup) were calculated using the paired *t* test to compare baseline values versus values at week 12 within each group. \* indicates statistically significant differences (*p* < 0.05). Abbreviations: WBC, white blood cell; RBC, red blood cell; HGB, hemoglobin; HCT, hematocrit; MCV, mean corpuscular volume; MCH, mean corpuscular hemoglobin; MCHC, mean corpuscular hemoglobin concentration; RDW-CV, red cell distribution width-coefficient of variation; AST, aspartate aminotransferase; ALT, alanine aminotransferase; γ-GT, gamma-glutamyl transferase; BUN, blood urea nitrogen; MDRD-based eGFR, modification of diet in renal disease equation-based estimated glomerular filtration rate.

## Discussion

In this randomized controlled trial, we investigated the effects of *L. plantarum* LP28 supplementation on upper respiratory symptoms in middle-aged and older adults over a 12-week period. Although no statistically significant group-by-time interaction effects were observed for most outcomes, several directional trends were evident. The clinical symptom outcomes among participants who developed colds showed mixed patterns. Among infected participants, compared with those in the placebo group, those in the probiotic group presented lower peak discomfort scores, average daily discomfort scores, and AUC values; however, none of these differences reached statistical significance (**Table 2**). These findings indicated only a modest tendency toward reduced symptom burden with LP28 supplementation. Interestingly, an age-stratified subgroup analysis further revealed distinct clinical effects of LP28 supplementation between middle-aged (< 65 years) and older (≥65 years) individuals. In symptomatic participants aged under 65 years **(Supplementary Table 1)**, LP28 supplementation had a symptom-mitigating effect, although no significant difference in the incidence of the common cold was observed between the LP28 and placebo groups [48.00% (12/25) vs. 46.88% (15/32); Fisher’s exact test, *p* = 1.000]. Compared with the placebo group, the LP28 group had a lower average discomfort score (5.47 ± 2.74 vs. 10.59 ± 6.76, *p* = 0.077), peak intensity (9.78 ± 6.08 vs. 31.08 ± 35.48, *p* = 0.025), and AUC for overall symptoms (37.56 ± 60.82 vs. 112.73 ± 140.16, *p* = 0.025), with substantial effect sizes (Cliff’s δ: -0.35 to -0.58). Conversely, no significant alleviation of symptoms was observed among symptomatic adults aged ≥ 65 years **(Supplementary Table 2)**. However, compared with placebo, LP28 supplementation significantly reduced the incidence of the common cold [19.05% (4/21) vs. 61.11% (11/18); Fisher’s exact test, *p* = 0.010]. Together, these data demonstrated an age-dependent difference: LP28 provided primary prophylaxis against infection in older adults (≥ 65 years) but effectively attenuated acute symptom severity in middle-aged individuals (< 65 years). With respect to upper respiratory symptoms, compared with the placebo group, the probiotic group demonstrated a consistently lower symptom burden and incidence across several domains, including cough, sneezing, runny nose, plugged nose and headache. However, these differences did not reach statistical significance. These findings are consistent with those of previous randomized controlled trials and meta-analyses indicating that probiotics may reduce the incidence of upper respiratory tract infections (URTIs) without exerting strong or consistent effects on symptom severity once infection occurs [7–9]. For example, Zhang et al. reported that probiotic yogurt supplementation significantly reduced the risk of URTIs in adults with frequent colds [8], whereas Pu et al. reported a lower URTI risk in healthy elderly individuals who consumed probiotic yogurt than in those who did not receive supplementation [7]. Similarly, Cochrane reviews concluded that probiotics are more effective in preventing acute URTIs and reducing episode frequency and duration than in producing large changes in symptom intensity [9]. Several factors may explain the absence of statistically significant findings in the present study. First, the participants were generally healthy individuals with relatively mild baseline symptoms, limiting the potential for measurable improvement. Second, variability in self-reported symptom data may have reduced statistical power. Third, the sample size may not have been sufficient to detect small effect sizes, particularly after adjustment for multiple comparisons. Furthermore, the health benefits of probiotics are strain-specific and strongly influenced by interindividual host variations [7–10]. Differences in baseline microbiome composition and host metabolic or immune status can alter individual responsiveness to probiotic therapy. Such host heterogeneity represents a key factor that may attenuate measurable differences between groups. Despite these limitations, subtle trends were observed in upper respiratory outcomes, suggesting a potential overall effect of probiotic supplementation. Rather than exerting strong effects on individual symptoms, probiotics may contribute to modest but broad regulatory effects on host health through immune modulation and maintenance of gut homeostasis. Consequently, probiotic interventions may function primarily through preventive and immunomodulatory pathways rather than direct symptom mitigation.

As critical cellular components of innate immunosurveillance, NK cells play an essential dual role in host defense: rapidly eliminating virus-infected cells and selectively destroying tumor cells. A central finding of this study is the enhancement of the innate immune response, characterized by a significant increase in NK cell cytotoxicity without a compensatory expansion of total lymphocyte populations (**Figure 3**). Importantly, compared with that in the placebo group, NK cell activity in the LP28 supplementation group was significantly elevated but reached a steady physiological plateau rather than showing a continuous, progressive increase over time. This controlled activity profile confirms that *L. plantarum* LP28 acts as a homeostatic “functional primer” that optimizes baseline cytotoxic capacity, thereby ruling out concerns of immune hyperactivation or sustained inflammatory overstimulation. Significantly, preserved NK cell cytotoxic activity, rather than expansion of the total lymphocyte population, is a well-established immunological hallmark observed in healthy centenarians and longevity cohorts and directly contributes to sustained immune resilience during late life [11]. The ability of *L. plantarum* LP28 to specifically restore and sustain NK cell effector function without inducing lymphoproliferative expansion strongly aligns with these longevity phenotypes. Therefore, restoring and increasing NK cell cytotoxicity represents a crucial clinical strategy to counteract age-related immune dysfunction [12, 13]. By “activating” these primary effector cells, the probiotic enables more rapid and efficient clearance of viral pathogens at the mucosal entry point, directly accounting for the decreased URTI frequency observed in the probiotic cohort [14, 15].

The mechanism underlying *L. plantarum* LP28-induced immunomodulation may involve the targeting of the IL-12 pathway. Our cytokine results revealed a significant increase in serum IL-12 levels in the probiotic group at week 6 (**Figure 5C**). As IL-12 is a master regulator of Th1 immunity and a potential activator of NK cells, elevation of its concentration provides a clear biochemical rationale for the observed increase in cytotoxic activity [16, 17]. Remarkably, this functional “priming” was achieved without inducing a chronic inflammatory state [18]. Age-associated immune dysregulation is frequently accompanied by “inflammaging”—a state of systemic, low-grade chronic inflammation characterized by elevated baseline proinflammatory cytokine levels that accelerate cellular senescence and susceptibility to URTIs [19]. In contrast, healthy aged individuals maintain tightly controlled, stable inflammatory profiles. The stability of serum IFN-γ, IL-6, and TNF-α levels combined with the significant reduction in CRP levels underscore the effect of the probiotic as a “smart” immunomodulator. It strengthens the host against viral infection while simultaneously mitigating inflammation. Furthermore, the improvement in GI function induced by the probiotic may reflect successful establishment of the gut–immune axis. The probiotic probably interacts with gut-associated lymphoid tissue to promote these systemic immune effects, while salivary sIgA and α-amylase levels remained stable in the probiotic group. The levels of these markers indicated that the probiotic had a stabilizing effect on mucosal homeostasis by preventing the transiently decreasing in α-amylase levels observed in the placebo group.

Importantly, safety evaluation demonstrated that all hematological and biochemical parameters remained within normal physiological ranges throughout the 12-week supplementation period. The decrease in WBC count following LP28 supplementation may have been associated with the attenuation of chronic low-grade inflammation; it has been reported that certain probiotic strains exert such an effect through the modulation of gut– immune interactions and systemic immune responses [20, 21]. Since all hematological values remained within normal reference ranges, the change was unlikely to represent adverse hematological suppression. Interestingly, significant fluctuations in the amount of MCH were detected in both groups. Notably, hemoglobin levels and the mean corpuscular volume (MCV) remained stable throughout the study period. This pattern suggests that the observed variation in the amount of MCH is unlikely to represent a clinically meaningful change in erythropoiesis or iron metabolism but rather reflects normal biological and analytical variability. Given that all hematological indices remained within physiological ranges, these changes were not considered clinically significant. In addition, no evidence of hepatic or renal toxicity was observed, as reflected by stable AST, ALT, γ-GT, BUN, and creatinine levels and a stable eGFR. These findings support the overall safety and tolerability of daily LP28 supplementation.

In conclusion, the probiotic strain *L. plantarum* LP28 exerts potent and complex immunomodulatory effects. The increase in the activity of the IL-12/NK cell axis was specifically evidenced by significant increases in NK cell cytotoxicity and IL-12 levels and the maintenance of systemic inflammatory balance through a reduction in the level of CRP. *L. plantarum* LP28 provides robust defense against respiratory infections. This is clinically reflected in the notable reductions in cold incidence and cumulative symptom burden among middle-aged and older adults. Given that heightened infection susceptibility is a hallmark of age-related immunosenescence, the ability of *L. plantarum* LP28 to effectively lower URTI occurrence, increase the functional capacity of NK cells, and optimize inflammatory profiles underscores its clinical value in promoting healthy aging. Collectively, these findings suggest that *L. plantarum* LP28 supplementation is a comprehensive and safe nutritional strategy to enhance innate immunity and maintain physiological homeostasis, effectively counteracting age-related immune decline.

## Acknowledgments

The technical support provided by the Instrument Center of the Department of Medical Research of Taichung Veterans General Hospital is gratefully acknowledged. We also thank SYNBIO TECH INC. Kaohsiung City, Taiwan, for providing the probiotics.

## Author Contributions

J.-J.S., C.-L.W., H.-Y.H. and C.-C.L. designed the study. Z.-Y.L., M.-C.C. and M.-H.T. carried out the experiments. H.-Y.H., C.-C.L. and H.-W.H. provided reagents and other laboratory supplies. Z.-Y.L., M.-H.T., C.-M.S., H.-Y.H. and C.-C.L. analyzed the data. J.-J.S., C.-L.W., Z.-Y.L., M.-H.T., C.-M.S., H.-Y.H. and C.-C.L. prepared figures and wrote the manuscript. All authors have read and agreed to the published version of the manuscript.

## Funding

This study was funded by projects from the University–Industry Cooperation Fund, National Chung-Hsing University (113D530).

## Institutional Review Board Statement

The study was conducted according to the guidelines of the Declaration of Helsinki and approved by the Institutional Review Board of Taichung Veterans General Hospital (Taichung City, Taiwan; No. SF24108C).

## Informed Content Statement

Informed consent was obtained from all subjects involved in the study. Before the experiment, the researchers explained the experimental process in detail, and the experiment began after the subjects signed the consent form.

## Data Availability Statement

The data presented in this study are available on request from the corresponding author. The data are not publicly available due to the test sample patent and subject’s privacy and confidentiality.

## Conflicts of Interest

H.-Y.H., C.-C.L. and H.-W.H. are employed by SYNBIO TECH INC. J.-J.S., C.-L.W., Z.-Y.L., M.-H.T., C.-M.S. and M.-C.C. declare no conflicts of interest.

## Notes

### Clinical Trial

NCT06724185

### Author Declarations

Ethical approval for this work was given by the Institutional Review Board of Taichung Veterans General Hospital (Protocol No. SF24108C).

