## Supplemental Table 1 for "The Influence of Long-Term Supplementation with *Lactiplantibacillus plantarum* LP28 on NK Cell Activity and Upper Respiratory Symptoms in Middle-Aged and Older Adults: A Randomized, Double-Blind, Placebo-Controlled Trial"

**Supplementary table 1. Assessment of URTI symptom severity in symptomatic participants aged under 65 years according to the WURSS.**

| | Probiotic group | Placebo group | *p* value | Effect size (Cliff's δ) |
|---|---|---|---|---|
| Cold occurrence (%) | 48.00% (12/25) | 46.88% (15/32) | 1.000 | OR = 0.96 (95% CI: 0.30-3.10) |
| Among participants with cold (infection-only analysis) | | | | |
| Number of discomfort days | 9.22 ± 12.92 | 13.46 ± 19.83 | 0.178 | -0.35 |
| Discomfort score (AVG) | 5.47 ± 2.74 | 10.59 ± 6.76 | 0.077 | -0.46 |
| Discomfort score (peak) | 9.78 ± 6.08 | 31.08 ± 35.48 | 0.025* | -0.58 |
| Discomfort score (AUC) | 37.56 ± 60.82 | 112.73 ± 140.16 | 0.025* | -0.58 |

The data are presented as mean ± SD. Between-group comparisons of cold occurrence were performed using Fisher's exact test, with the odds ratio (placebo vs. probiotics) estimated by conditional maximum likelihood (95% CI). Symptom parameters among participants with a cold (infection-only analysis) were compared using the Mann–Whitney *U* test, and effect sizes were estimated using Cliff's δ (negative values favor the Probiotic group). Infection-only analyses included participants in the probiotic ($n$ = 9) and placebo ($n$ = 13) groups who reported a cold and who completed the WURSS. * $p < 0.05$.
