## Supplemental Table 2 for "The Influence of Long-Term Supplementation with *Lactiplantibacillus plantarum* LP28 on NK Cell Activity and Upper Respiratory Symptoms in Middle-Aged and Older Adults: A Randomized, Double-Blind, Placebo-Controlled Trial"

**Supplementary table 2. URTI symptom metrics derived from the WURSS for symptomati older participants (≥65 years).**

| | Probiotic group | Placebo group | $p$ value | Effect size (Cliff's δ) |
|---|---|---|---|---|
| Cold occurrence (%) | 19.05% (4/21) | 61.11% (11/18) | 0.010* | OR = 6.31 (95% CI: 1.31-37.46) |
| Among participants with cold (infection-only analysis) | | | | |
| Number of discomfort days | 6.33 ± 5.51 | 13.88 ± 29.28 | 0.757 | 0.17 |
| Discomfort score (AVG) | 17.92 ± 16.99 | 7.21 ± 8.01 | 0.413 | 0.38 |
| Discomfort score (peak) | 29.33 ± 30.01 | 16.38 ± 27.18 | 0.608 | 0.25 |
| Discomfort score (AUC) | 175.33 ± 230.38 | 75.31 ± 116.29 | 0.610 | 0.25 |

The data are presented as mean ± SD. Between-group comparisons of cold occurrence were performed using Fisher's exact test, with the odds ratio (placebo vs. probiotics) estimated by conditional maximum likelihood (95% CI). Symptom parameters among participants with a cold (infection-only analysis) were compared using the Mann–Whitney U test, and effect sizes were estimated using Cliff's δ (negative values favor the Probiotic group). Infection-only analyses included participants in the probiotic ($n$ = 3) and placebo ($n$ = 8) group who reported a cold and who completed the WURSS. * $p < 0.05$.
